# Genomic surveillance of SARS-CoV-2 during the first year of the pandemic in the Bronx enabled clinical and epidemiological inference

**DOI:** 10.1101/2021.02.08.21250641

**Authors:** J. Maximilian Fels, Saad Khan, Ryan Forster, Karin A. Skalina, Surksha Sirichand, Amy S. Fox, Aviv Bergman, William B. Mitchell, Lucia R. Wolgast, Wendy Szymczak, Robert H. Bortz, M. Eugenia Dieterle, Catalina Florez, Denise Haslwanter, Rohit K. Jangra, Ethan Laudermilch, Ariel S. Wirchnianski, Jason Barnhill, David L. Goldman, Hnin Khine, D. Yitzchak Goldstein, Johanna P. Daily, Kartik Chandran, Libusha Kelly

## Abstract

The Bronx was an early epicenter of the COVID-19 pandemic in the USA. We conducted temporal genomic surveillance of SARS-CoV-2 genomes across the Bronx from March-October 2020. Although the local structure of SARS-CoV-2 lineages mirrored those of New York City and New York State, temporal sampling revealed a dynamic and changing landscape of SARS-CoV-2 genomic diversity. Mapping the trajectories of variants, we found that while some became ‘endemic’ to the Bronx, other, novel variants rose in prevalence in the late summer/early fall. Geographically resolved genomes enabled us to distinguish between cases of reinfection and persistent infection in two pediatric patients. We propose that limited, targeted, temporal genomic surveillance has clinical and epidemiological utility in managing the ongoing COVID pandemic.

**One sentence summary:** Temporally and geographically resolved sequencing of SARS-CoV-2 genotypes enabled surveillance of novel genotypes, identification of endemic viral variants, and clinical inferences, in the first wave of the COVID-19 pandemic in the Bronx.

## Main text

COVID-19 continues to have a devastating effect on the health of communities across the globe, with over 79 million reported cases and greater than 1.7 million deaths since the start of the pandemic as reported on December 29th, 2020 (*1*). The Bronx, a borough of New York City (NYC) sustained the second highest rate of COVID-19 in early waves of the pandemic in New York City with 6,035 cases per 100,000 people as of January 11, 2021 (*2*). To track the local spread of SARS-CoV-2, we conducted a genomic epidemiologic study at Montefiore Health Systems (MHS), which offers healthcare services to two million residents throughout the Bronx, one of the most diverse and poorest urban communities in the United States.

The number of COVID-19 cases peaked in the Bronx in March–April 2020 and subsided during the late spring into summer 2020. To characterize the genetic diversity of SARS-CoV-2 in the Bronx, we randomly selected nasopharyngeal remnant clinical samples positive for SARS-CoV-2 by RT-PCR testing from the MHS clinical laboratory between March and October 2020. Genomic viral RNA was extracted from nasopharyngeal swabs, and sequencing libraries were prepared using the ARTIC Network protocol and analyzed on an Oxford Nanopore MinION (*3, 4*). The ARTIC Network bioinformatics protocol was used to quality check and annotate SARS-CoV-2 genomes with default parameterization (*5*). We called variants with the NextClade tool and annotated lineages using the October 13th, 2021 update of PANGOLIN version 3.1.14 (*6, 7*). Samples were derived from patients who required hospitalization (48%), mild disease managed as outpatients (26%) and asymptomatic carriers (8.9%) (**Fig. 1A**).

**Figure 1.**
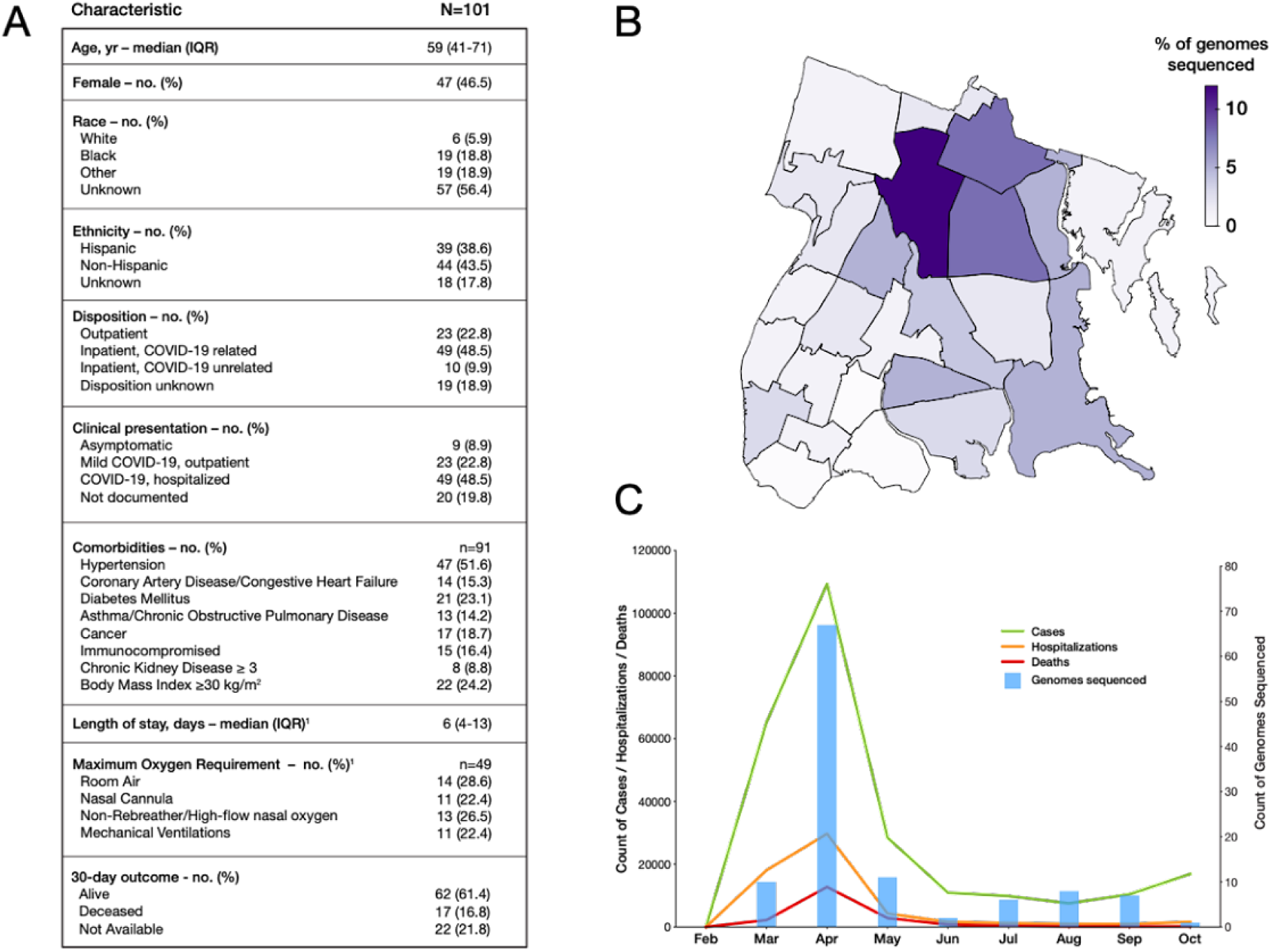
Surveilling SARS-CoV-2 genomes in the Bronx. **A)** Table of clinical characteristics of sampled patients. **B)** SARS-CoV-2 genomes sequenced per Zip code in NYC. Darker colors indicate heavier sampling; **C)** SARS-CoV-2 genomes sequenced over time during the COVID-19 pandemic. Date is indicated on the x axis. Blue bars and the associated right hand y axis indicate the number of genomes sequenced. The left-hand y axis represents different features of COVID-19 in the Bronx; green lines indicate COVID-19 cases, the red line deaths associated with COVID-19 and the orange line hospitalizations associated with COVID-19 in the Bronx.

We collected 137 samples, and from these generated 104 high-quality genomes from 101 patients with >95% coverage (**Fig. S1 and S2)**. Sequence data were derived from residents throughout the Bronx and were associated with 22 of 25 zip codes (**Fig. 1B)**. Genomic sampling was greatest at the onset of the COVID-19 pandemic in March and April, but intermittent sampling continued as caseloads declined over the summer and fall (**Fig. 1C**).

Analysis of the resulting 104 SARS-CoV-2 genome sequences revealed that the B.1 lineage was the most prevalent during the early months of the pandemic in the Bronx; however, several other lineages were also present at low frequencies throughout the sampling period (**Fig. 2A**). Many low-frequency B.1 sub-lineages were sampled at different time points. Some of these lineages were first observed elsewhere before being observed at MHS. Two lineages, B.1.604 and B.1.448, were first observed in the MHS and subsequently spread to other areas of the United States. We sampled the first 5 of 48 B.1.604 assigned genomes, for which spread was limited to the Northeastern United States. B.1.448 represented a larger sub-lineage of B.1 that appeared to arise from the Bronx, having 332 other genomes sharing lineage defining SNPs. Both of these lineages were sampled until early 2021, and B.1.604 was only sampled at the MHS during the time frame of March to October. The majority of Bronx genomes were classified as sub-lineages of B.1 or B.1.1; the B.1 lineage continued to be sampled until late August. From March to October the Bronx SARS-CoV-2 lineages represent a subset of SARS-CoV-2 lineage diversity present in NYC, New York State (NYS), the United States (US) and the world (**Fig. 2B**) (*8, 9*). The prevalence of Bronx originating lineages B.1.448 and B.1.604 did not change when comparing the Bronx to NYS. B.1.448 was found in other areas of the US and world during the sampling period, whereas B.1.604 was only observed in the Bronx. We noted that most B.1 and B.1.1 sub-lineages were low in number compared to the B.1 and B.1.1 lineages, while the B.1.1 relative representation at MHS, NY and US was higher than the global representation. To determine how the Bronx sequences of these lineages compared to those sampled across the world, we created a downsampled SARS-CoV-2 tree from 613 high-quality SARS-CoV-2 genomes deposited in GISAID with available location and collection dates from March to October 2020. We found that Bronx SARS-CoV-2 sequences represented subsets of different clades of the global tree (**Fig. 2C**).

**Figure 2.**
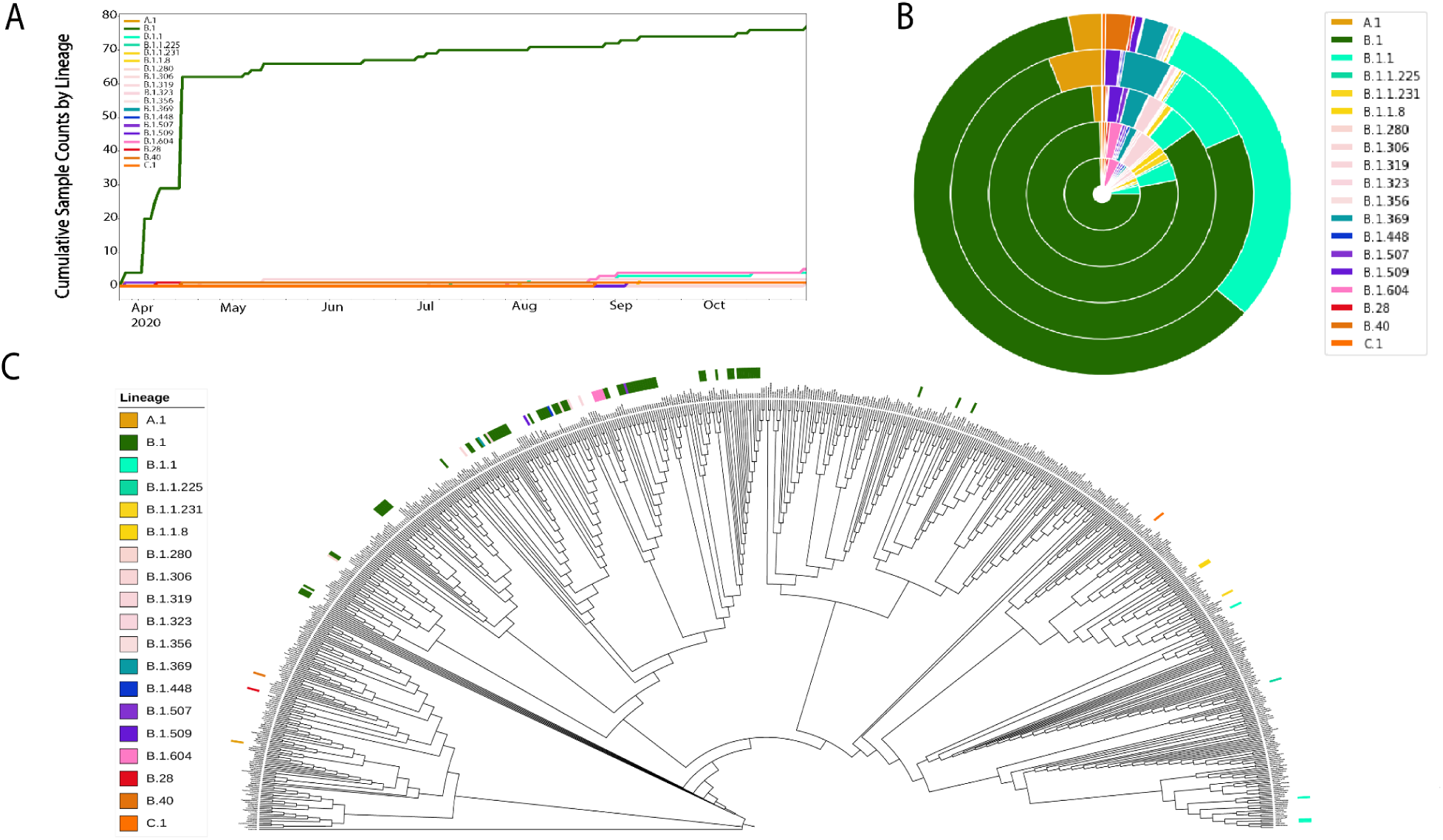
Bronx SARS-CoV-2 genome lineages in the context of local and global sampling. **A)** Cumulative counts of PANGOLIN guide tree-based lineage assignments plotted against time; **B)** Prevalence of lineages seen in the Bronx compared to their prevalence in other regions. Inner to outer rings represent the Bronx, New York State, USA, the world, respectively. Lineage coloring is the same as A); **C)** Phylogeny of the Bronx strains in the context of SARS-CoV-2 strains from around the world. Bronx strains and their associated lineages are indicated with colored lines.

We next examined patterns in variant nucleotide positions observed in our data. We found that variation is distributed across the SARS-CoV-2 genome and that some variants are present in almost all Bronx genomes sequenced—these can be described as ‘core’ to the Bronx at present (**Fig. 3A**). Core variants include the spike protein variant A23403G (D614G), as well as variants C241T, C1059T (T265I), C3037T, C14408T (P314L) in Orf1ab, and G25563T (Q57H) in Orf3a. We next examined the dynamics of individual SARS-CoV-2 variants. Although the core variants continued to increase in prevalence as we sequenced new genomes, we also observed variants novel to the Bronx whose prevalence increased, whereas others plateaued (**Fig. 3B**).

**Figure 3.**
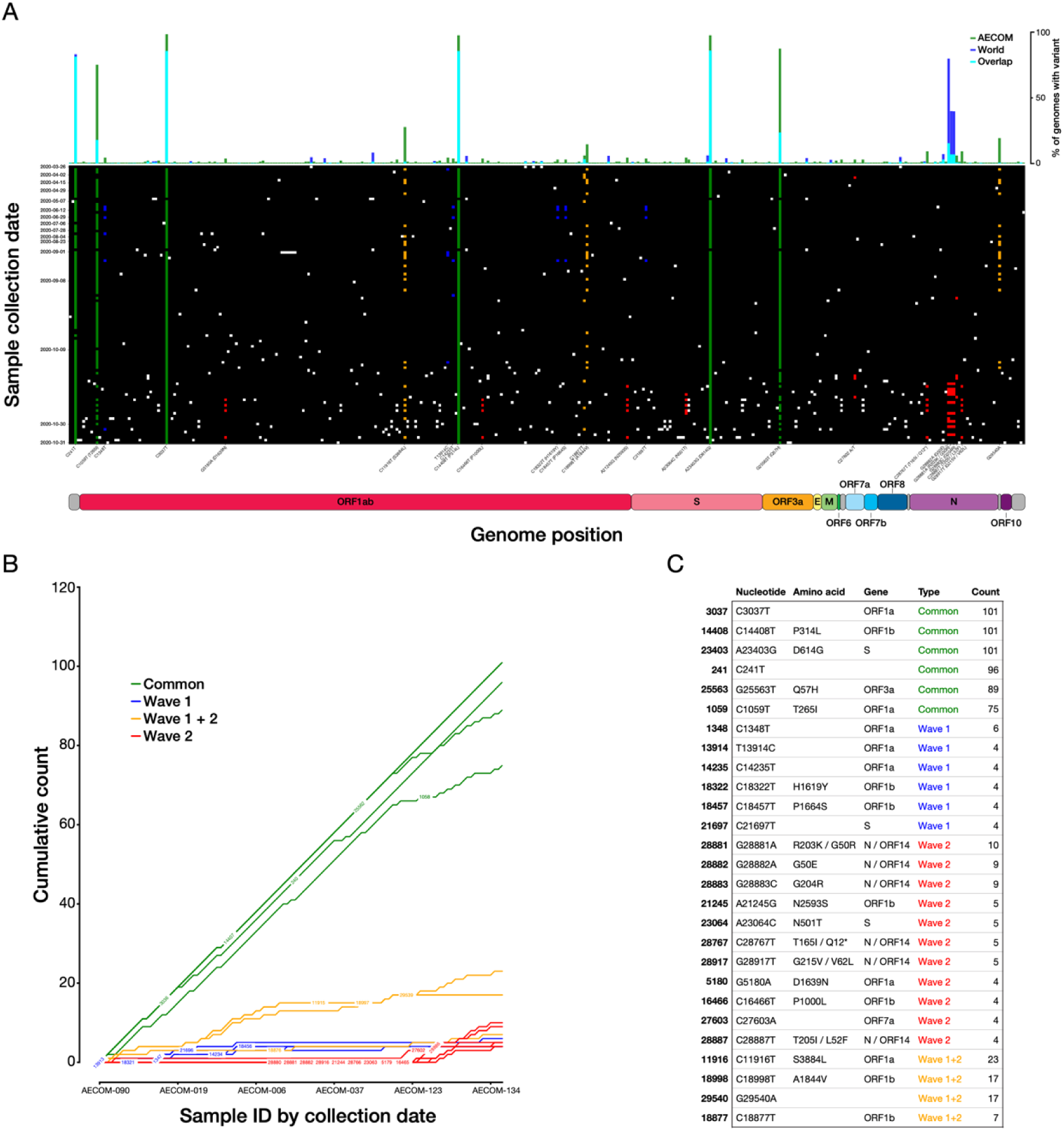
SARS-CoV-2 variants and their trajectories in the Bronx. **A)** Individual SARS-CoV-2 variants plotted across the viral genome (x axis), with genomes sorted by sampling date (y axis). Positions that are variable with respect to the reference SARS-CoV-2 strain are shown with a white (low-frequency), green (common), yellow (wave 1+2), blue (wave 1) or red (wave 2) squares. The histogram across the top plots the prevalence of a given variant across all Bronx SARS-CoV-2 genomes in this study relative to the world; **B)** Rarefaction curve of cumulative variant counts over time for variants observed at least four times in the Bronx SARS-CoV-2 genomes set; **C)** Table showing details for variants in 3B.

In the spike protein, we found amino acid variants D614G (core), N501T in 5 patients, and both N501Y and P681R in one patient. We noted that P314L in Orf1b is also a core variant in our dataset, reflecting observations in other studies that this variant is in linkage disequilibrium with D614G (*10*). We did not observe the B.1.1.7 lineage first identified in the United Kingdom in the fall of 2020, or any other WHO classified strains that were circulating during this time frame, known to contain the N501Y variant and similarly the P681H in our samples. The N501 residue of the spike protein is part of the receptor binding domain and the receptor binding motif, and variants at this position may influence ACE2 receptor binding (*11*). In comparing Bronx variants to those found in the rest of the world, we found that some variants, such as the spike protein D614G variant, are prevalent both in our set and in the world; however, some ‘core’ Bronx variants such as C1059T (T265I in Orf1ab) and G25563T (Q57H in Orf3a) are not as prevalent in the rest of the world at this time period **(top bar Fig. 3A, Fig. 3C, Fig S3)**. The geographic specificity of variants creates a fingerprint that can be useful for tracing the spread of particular variants; a lineage containing the variant C2416T, linked to the Boston Biogen COVID-19 outbreak, could be traced to infections around the world (*12*). The C2416T variant was also observed in three patients in our dataset. We note that rare variants are uniformly distributed throughout the sampling period (**Fig. S4**) and further that the functional impact of these variants is not well resolved.

A phylogenetic tree of SARS-CoV-2 shows that strains collected earlier in the pandemic are distinguishable from strains collected later, suggesting that new strains were being continuously introduced into the Bronx (**Fig. 4**, inner ring, red indicates earlier samples, green newer samples). In considering the lineages of SARS-CoV-2, there was evidence of ongoing presence of B.1 lineage throughout the study period, starting from the onset of pandemic until the end of the study period (**Fig. 4**, outer ring indicates lineage). We found that the B.1 sub-lineages, such as the parent lineage of variants of concern alpha and omicron: B.1.1 and Bronx originating lineages: B.1.448 and B.1.604, had increasing presence in the latter part of the study period and that newer collections of B.1 strains, which cluster away from older B.1 sequences, appear at later sampling dates. There are newer B.1 and B.1 sub-lineages that form a distinct clade from older B.1 lineages in the Bronx SARS-CoV-2 tree. We posit that these two clades reflect two different types of SARS-CoV-2 isolates: those that were circulating locally and those that were newly introduced. We considered SARS-CoV-2 isolates that group on the downsampled global tree and group on the local Bronx tree with our first wave pandemic sampling to be ‘circulating.’ We observed that the Bronx originating lineages pair with what we consider circulating strains of B.1 on the global tree. However, we continue to observe isolates that fall into this ‘first wave’ clade of B.1 during the summer, post-first wave, and therefore consider these to have persisted in the Bronx. We consider ‘introduced’ isolates those that are newer sequences in the local Bronx tree that are also spread out in different clades across the global tree, or arising from lineages that originate outside the Bronx. This approach is useful for identifying isolates that may be precursors to new sublineages (**Fig. 2C and 4)**.

**Figure 4.**
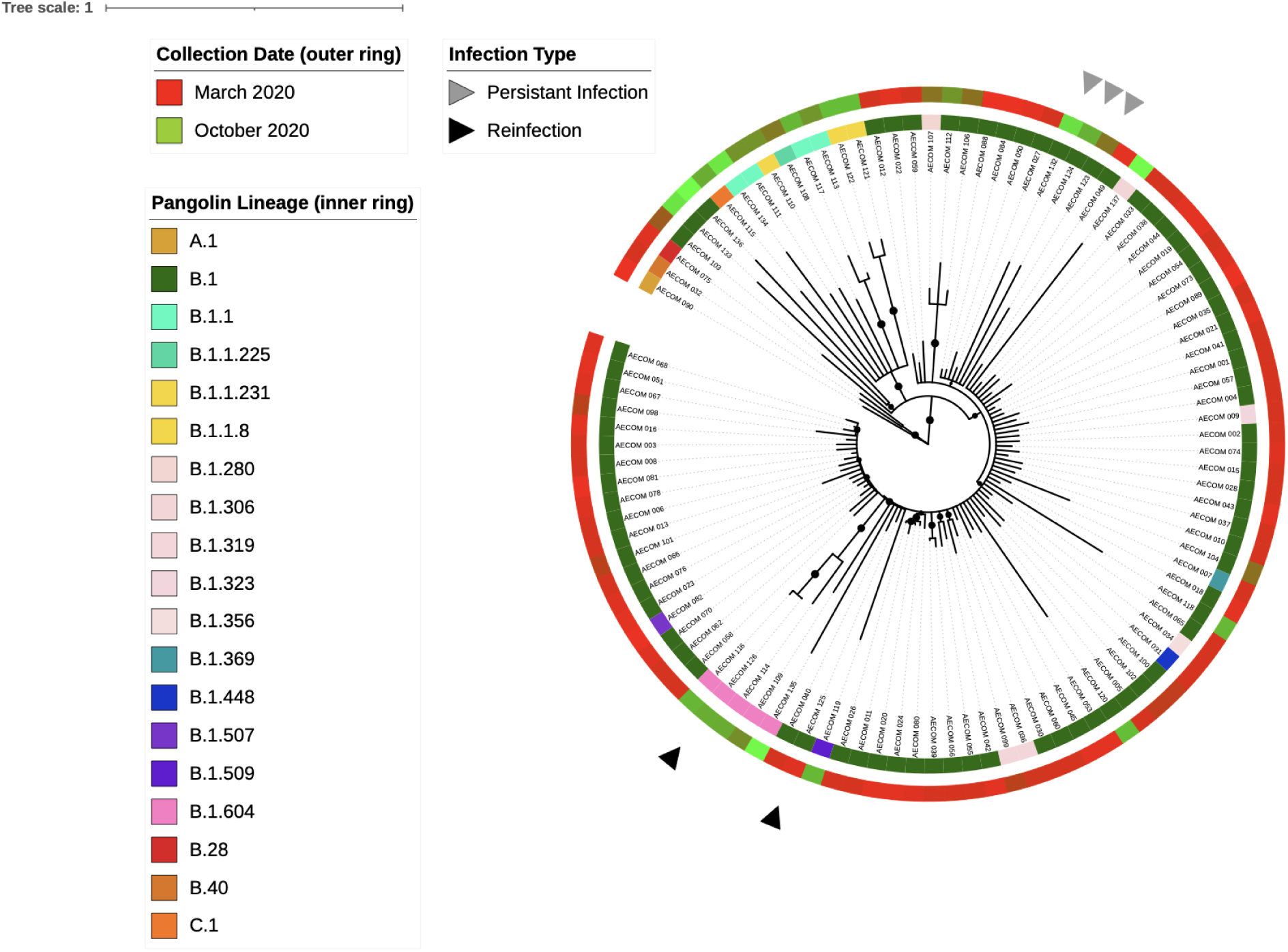
Clinical relevance of the changing genomic landscape of SARS-CoV-2 in the Bronx. Phylogenetic tree based on whole genome alignments of Bronx isolates. Colored rings around the tree indicate SARS-Cov-2 lineage (inner ring) and the date of sampling (outer ring, red=earlier, green=later). Samples from the same patient are indicated with symbols; a reinfection case is indicated with black arrows and a putative persistent infection case is indicated with grey arrows. Black circles on the branches indicate bootstrap values of 85 or greater. Tree was generated with TimeTree and visualized with iTOL (*13, 21*).

This local phylogenetic framework of SARS-CoV-2 strains in the Bronx enabled us to distinguish between a case of reinfection and a case of persistent infection in two pediatric patients. The first case is a 12 to 18-y.o. female who was initially seen in April 2020 in the emergency department with 3 days of fever, sore throat, anosmia, and ageusia in the setting of the death of her father at home from suspected COVID-19. SARS-CoV-2 infection in this patient was confirmed by RT-PCR. She had a total of 6 days of symptoms and was in general good health until the second presentation. In August 2020, she presented again to the emergency department with two days of fever, severe postprandial abdominal cramps, watery diarrhea and generalized body aches. All other reviews of symptoms were negative. The patient had no known COVID-19 exposures and limited outside exposure with visits only to supermarkets and parks near her home. A respiratory pathogen panel was negative but her SARS-CoV-2 RT-PCR was positive, as was her SARS-COV-2 IgM Immune Status Ratio (ISR) (2.1, with less than 1 considered negative). Her IgG ISR was negative, 8.7 (normal range ISR < 9). The patient had a total of three days of fever with complete resolution of all other symptoms by day four of illness.

The two SARS-CoV-2 genomes sequenced from this patient were 142 days apart and differed in nucleotide sequence at 17 different positions. The first and second samples from this patient have different lineage designations and fall in different local phylogenetic clades in the Bronx phylogenetic tree, supporting the hypothesis that we are observing a new infection and not prolonged shedding from the original SARS-CoV-2 infection. In fact, the patient was reinfected with one of the first few isolates of the Bronx-originating lineage: B.1.604, suggesting they belong to a separate transmission chain (**Fig. 4, black arrows**). Given the history of limited exposures to high-risk activities for this patient between the two episodes and the overall low incidence of SARS-CoV-2 infection in New York at the time of the second presentation in August, genomic and phylogenetic analysis provided key confirmatory evidence in support of the clinical inference of a reinfection.

The second case involved a 12 to 18-y.o. female with an incompletely characterized immunodeficiency who presented in July 2020 with an oral lesion. She had no fever, or respiratory or gastrointestinal symptoms, and had neutropenia (absolute neutrophil count 700 cells/ul). After admission for further evaluation, she was found to be SARS-CoV-2 positive. During the admission, she was intermittently febrile and neutropenic and was treated with broad spectrum antibiotics. She developed a buttock lesion that was biopsied, revealing a thrombotic vasculopathy with infarction. Due to concern that the lesion could represent COVID-19–associated vasculopathy, and in the setting of persistent fever and intermittent neutropenia, she was treated with a 10-day course of remdesivir. The patient continued to have positive nasopharyngeal swabs for SARS-CoV-2 from early July to the end of September (**Table S1 and Fig. S5**). Her SARS-CoV-2 IgG (Abbott) was negative in mid-August.

For this patient, the three sequenced SARS-CoV-2 genomes sampled in July, August and October are members of the B.1 lineage and fall in the same clade (**Fig. 4, grey arrows**). This clade is polytomic by TimeTree, meaning that it is not possible to resolve the relationships between sequences within this clade, but the clade itself is supported by a bootstrap value of 870/1000 (SH-aLRT replicates) (*13, 14*). We therefore posit that the three strains sequenced from this patient, despite having some variation, are more likely to represent a single SARS-CoV-2 infection rather than multiple infections. Together, these genomic, phylogenetic, and clinical observations strongly suggest that this patient has been unable to clear a single infection of SARS-CoV-2, as opposed to being reinfected with a distinct strain. Other examples of persistent infection with SARS-CoV-2 have been reported, but not, to our knowledge, in children (*15–17*). A woman diagnosed with chronic lymphocytic leukemia who was sampled 5 times, had SARS-CoV-2 sequences displaying intrahost variation despite the SARS-CoV-2 being polytomic, similar to what we observe here (*18*). The polytomy that encompasses this persistent case also contains independent local strains of SARS-CoV-2 that do not separate on the global tree, suggesting that some variants seen in this patient are also shared locally in the Bronx (**Fig. 2C and 4**).

Our work supports guiding principles for practical and clinical applications of SARS-CoV-2 sequencing in the COVID-19 pandemic. How many genomes do you need to sequence for a local community to resolve clinical questions? In our case, ∼100 genomes were sufficient to place new patients into the context of the variability of SARS-CoV-2 during the pandemic and to be able to answer coarse-grained questions to determine reinfection vs. persistent infection and community-level observations of older vs. newly introduced strains. The targeted utilization of small numbers of stored swabs for temporally resolved viral genomic surveillance could thus resolve clinical questions related to persistence vs. reinfection. This localized molecular and temporal description of SARS-CoV-2 genomes during the first wave of the COVID-19 pandemic in the Bronx, NY demonstrates the value of targeted, local sequencing efforts to guide clinical inference, and serves as a valuable baseline for ongoing studies of waves of the pandemic in this underserved urban community.

## Data Availability

All sequences generated in this study have been made publicly available through the GISAID hCoV-19 sequence database. The source code used for sequencing, analysis, and figure generation, is hosted on Github at https://github.com/kellylab/genomic-surveillance-of-the-bronx.

## Acknowledgements

We thank Isabel Gutierrez, Estefania Valencia, and Laura Polanco for laboratory management and technical assistance and the Chandran and Kelly labs for helpful comments on the manuscript. We thank NextStrain, GISAID, and all labs who contributed SARS-CoV-2 sequences for public access. We thank the healthcare workers and patients of the Montefiore Healthcare System.

## Author contributions

Study design: LK, KC, JPD, DG, HK, DG. Data collection and analysis: SK, RF, JMF, LK, KC, AB. Clinical and experimental support: KAS, SS, ASF, WBM, LRW, RHBIII, MED, CF, DH, RKJ, EL, ASW, JB. Wrote paper: LK, KC, JMF, SK, RF. Edited paper: all authors.

## Supplementary materials

Materials and Methods

Figs. S1 to S5

Table S1

