## Supplemental and Methods for "Genomic surveillance of SARS-CoV-2 during the first year of the pandemic in the Bronx enabled clinical and epidemiological inference"

**This PDF file includes:**

Materials and Methods

Figs. S1 to S5

Table S1

### Material and methods

#### Ethics statement

Remnant nasopharyngeal swabs were collected and deidentified at Montefiore Medical Center. This work was approved by the Institutional Review Board of Albert Einstein College of Medicine under IRB number 2016-6137.

#### Data availability

All sequences generated in this study have been made publicly available through the GISAID hCoV-19 sequence database. The source code used for sequencing, analysis, and figure generation, is hosted on Github at https://github.com/kellylab/genomic-surveillance-of-the-bronx.

#### RNA isolation

Viral RNA was isolated from nasopharyngeal swabs using the MagMAX Viral RNA isolation kit (Applied Biosystems, #AM1939) according to the manufacturer's specification. 400 µl of viral transport medium was used as input for each sample. Isolated RNA was then stored at -80C prior to sequencing library generation.

#### Preparation of sequencing libraries

Sequencing libraries were prepared according to the protocol established by the ARTIC network (*3*, *4*). Briefly, cDNA was generated from viral RNA using SuperScript IV reverse transcriptase (Thermo Scientific, #18090010). 400 nt tiled amplicons were generated using the V3 primer pool, divided into 4 sub-pools for increased efficiency. Amplification was performed using Q5 High-Fidelity polymerase (New England Biolabs, #M0491S) with cycle numbers optimized for each sub-pool. Following amplicon cleanup using AMPure XP beads (Beckman Coulter, #A63880), 5 ng of input DNA, quantified using Quant-iT PicoGreen dsDNA Assay Kit (Invitrogen, #P7589), was natively barcoded using the Native Barcoding Expansion (Nanopore, # EXP-NBD104). After another round of amplicon cleanup using AMPure XP beads, sequencing adapters were ligated to pooled barcoded amplicons using NEBNext Quick Ligation Module (New England Biolabs, #E6056). Following an additional step of cleanup and quantification, the final libraries were sequenced.

**Nanopore MinION sequencing**

Sequencing libraries were diluted in elution buffer (Qiagen, # 19086) to a concentration corresponding to approximately 20 ng of library per sequencing run. MinION flow cells (Oxford Nanopore, #FLO-MIN106D were prepared using the Ligation Sequencing Kit (Oxford Nanopore, #SQK-LSK109). Libraries were then loaded onto the flow cell and sequencing allowed to proceed for 10 to 20 hours depending on library size.

**Sequencing analysis**

ONT MinION output files in fast5 format were processed using an implementation of the ARTIC sequencing pipeline on Google Cloud Platform. Briefly, this pipeline consists of the following steps: 1) Basecall reads using Oxford Nanopore’s Guppy tool; 2) Detect barcodes to sort out reads from different samples using Guppy; 3) Remove chimeric reads and small contaminations by filtering out all reads not within 400-700nt in length; 4) Align reads to the Wuhan reference genome (NCBI identifier MN908947.3) using minimap2, generate a consensus genome, and call variants using the nanopolish tool.

The pipeline was run using the workflow tool Argo running on a Kubernetes cluster in the cloud. Data was stored on a cloud storage bucket between steps (see supplemental code). Low-coverage sequences were improved by combining passed reads from multiple sequencing runs before generating consensus sequences.

**Quality control**

We included in our analysis only sequences that had 95% or higher coverage, a criterion 104 out of 132 sequences satisfied (**Fig S2)**. We also looked for signs of biases in the base calling pipeline which would result in higher or lower likelihood of gaps in certain regions. We found that the probability of a gap being present in the consensus is strongly correlated with the coverage level in the BAM file generated by the pipeline. In particular, we found that a coverage of 20x was almost always sufficient to result in a basecall being made at a given position but that the majority of positions had coverage above 400x. Thus, any biases in the pipeline are more likely to arise from biases in the nanopore sequencer itself or its basecaller rather than the consensus generation software.

**Variant annotation and global analysis of variants**

We used the NextClade command line tool to assign variant calls to each of the samples. This tool performs a pairwise alignment between an assembled genome and the Wuhan reference genome and reports the differences as variant calls. NextClade was also used to determine the amino acid changes implied by each variant. This method of variant calling was chosen over the one provided in the ARTIC pipeline in order to maintain consistency with our comparative analysis of global variant distributions.

We downloaded all of the 139676 genomes available from GISAID as of November 14,

2020 and used the NextClade command line tool to annotate each of their variants. This tool automatically rejects sequences that it deems of low quality, and this yielded variant calls from 139590 genomes from around the world. We used this output to compute the frequency of a variant as the percentage of samples in the world / AECOM dataset containing a given variant.

**Creating the local phylogenetic tree**

Individual FASTA files ≥ 95% coverage were collected after output by the ARTIC pipeline. The multi-FASTA was aligned using MAFFT on the Nextstrain command line interface version 1.16.7 (*6*, *22*). The resulting alignment FASTA generated was constructed into a maximum likelihood tree with 1000 SH-aLRT bootstraps using a TIM + F + I substitution model via iqtree-2.1.1-Windows (*14*). The tree was rooted on AECOM 90, the oldest outgroup sequence, and the entire tree was branch length corrected based on a fixed mutation rate of 0.0008 nucleotides/site/year with a standard deviation of 0.0004 using treetime 0.7.6 (*13*). The tree was visualized on iTOL and annotated with the iTOL annotation editor (*21*).

**Creating the global phylogenetic tree**

The GISAID database: GISAID - Initiative limited to 95% coverage and higher was used as an input for this analysis. The multi-FASTA of 11/14/2020 was filtered using the Nextstrain command line interface version 1.16.7 filter command. The specifications entailed and inclusion criteria used to construct a globally and temporally representative multi-FASTA was adapted from the criteria used to construct the Nextstrain global tree. An inclusion and exclusion text file was used to remove and keep strains that Nextstrain deemed important and is located here: https://github.com/nextstrain/ncov. The entire GISAID database was purged of any sequence with inconsistent metadata and grouped based on the country sequenced, the year, and month collected making 612 distinct groups from which 1 sequence was randomly chosen out of each group. The resultant multi-FASTA was aligned using MAFFT on the Nextstrain command line interface version 1.16.7 (*6*, *22*). A maximum likelihood tree was constructed with 1000 SH-aLRT bootstraps using a GTR substitution model via iqtree-2.1.1-Windows (*14*). The tree was visualized on iTOL and annotated with the iTOL annotation editor (*21*).

**Identifying lineages**

To identify pangolin lineages, the pangolin command line tool 2.0.8 was used in legacy mode, relying upon the 05/29/2020 update of the guide tree to assign lineages to local sequences via bootstrapping. The browse function of the GISAID database was used to count the lineages present in New York State. United States and global data were retrieved from SARS-CoV-2 lineages (cov-lineages.org) (*7*).

**Commands**

*Cat function and set encoding:*

Local: cat *.fasta > MSA_finale.txt

cat *txt > MSA_finale.fasta

Get-Content MSA_finale.fasta | Set-Content -Encoding utf8 MSA_finale_last.fasta

Global: cat *.fasta > Global_utf16.txt

cat *txt > Global_utf16.fasta

Get-Content Global_utf16.fasta | Set-Content -Encoding utf8 Global.fasta

*Nextstrain Augur Align:*

Local: augur align \

--sequences MSA_finale_last.fasta\

--reference-sequence config/ MN908947.3.gb \

--output aligned_MSA_finale_last.fasta \

--fill-gaps

--remove-reference

Global: augur align \

--sequences Global.fasta\

--reference-sequence config/MN908947.3.gb\

--output aligned_Global.fasta \

--fill-gaps

Output reads/ mafft specifications: using mafft to align via:

mafft --reorder --anysymbol --nomemsave --adjustdirection --thread 1 aligned_MSA_finale_last.fasta.to_align.fasta 1> aligned_MSA_finale_last.fasta 2> aligned_MSA_finale_last.fasta.log

Katoh et al, Nucleic Acid Research, vol 30, issue 14

https://doi.org/10.1093%2Fnar%2Fgkf436

*Iqtree:*

Local: bin\iqtree2 -s aligned_MSA_finale_last.fasta -m MFP -bb 10000 -alrt 1000

Global: bin\iqtree2 -s aligned_Global.fasta -m MFP -bb 10000 -alrt 1000 *treetime:*

treetime --tree MSA_finale_last.nwk  --dates Swab_samples_metadata.tsv --aln aligned_MSA_finale_last.fasta  --outdir Finale! --reroot AECOM_090 --clock-rate .0008 --clock-std-dev .0004 --keep-polytomies

*Nextstrain Augur Filter:*

```
augur filter \
  --sequences sequences.fasta \
  --metadata metadata.tsv \
  --exclude exclude.txt \
  --include include.txt\
  --output filtered.fasta \
  --group-by country year month \
  --min-length 27000 \
  --subsample-max-sequences 1000 \
  --exclude-where "division='USA' date='2020' date='2020-01-XX' date='2020-02-XX' date='2020-03-XX' date='2020-04-XX' date='2020-05-XX' date='2020-06-XX' date='2020-07-XX' date='2020-08-XX' date='2020-09-XX' date='2020-10-XX' date='2020-11-XX' date='2020-12-XX' date='2020-01' date='2020-02' date='2020-03' date='2020-04' date='2020-05' date='2020-06' date='2020-07' date='2020-08' date='2020-09' date='2020-10' date='2020-11' date='2020-12'"\
  --min-date 2019.74
```

*Pangolin Command Line Tool:*

pangolin MSA_finale_last.fasta --legacy --outfile Lineages.csv

**Figures and guide to scripts**

All figures were conjoined and post-processed to adjust colors and layout in Adobe Illustrator.

Figure 1

a) The table was generated by manually pulling EMR records for the patients whose samples came from the hospital. Not all of the fields were available for every patient.
b) The choropleth was generated by tabulating the zip codes for each of the patient samples which passed the 95% coverage threshold. We used the Python library Geopandas to then generate the choropleth using a map of the Bronx which we downloaded from NYC Open Data (https://data.beta.nyc/en/dataset/nyc-zip-code-tabulation-areas/resource/894e9162-871c-4552-a09c-c6915d8783fb).
c) This chart contains a histogram showing the distribution of samples in our dataset by month alongside three line plots showing the number of cases, hospitalizations, and deaths compiled by NYC Health (https://github.com/nychealth/coronavirus-data).

Figure 2

a) Chart of the cumulative density of lineages in our samples which passed the quality threshold as assigned using our Pangolin based method.
b) Data for this donut plot not derived from this study was acquired from SARS-CoV-2 lineages (cov-lineages.org).
c) Phylogenetic tree annotated by lineage using the online tool iTOL.

Figure 3

a) The top histogram shows the percentage of samples in the world / our dataset carrying a variant at a given position. For this histogram, we did not differentiate between different variants showing up at a position. The heatmap was generated by ordering our samples by date of sample collection and coloring in on the x-axis wherever a variant was found on a given position. Note that the x-axis is nonlinear in the sense that only positions where a variant was found in at least one sample in our cohort are included (this also holds true for the top histogram). We labeled on the x-axis all variants which showed up at least four times in our dataset and in parentheses indicated what the amino acid changes implied by those variants were. Some of the variants were in regions where multiple open reading frames overlap, so we indicated amino acid changes for both reading frames.

   For all three subfigures, we separated our variants into three categories:

   i) Rare: Positions where a variant is found less than four times.
   ii) Uncommon: Positions where a variant is found at least four times but less than 26 times (25% of our samples that were used in the analysis). This was further broken down into wave 1 and wave 2 categories, where a variant was considered to be 'wave-1-associated' if it showed up at least four times in the first half of the samples and at most once in the latter half (and vice-versa for wave 2). Samples that showed up at least once in both halves were labelled as wave 1 + 2.
   iii) Common: Positions where a variant is found at least 26 times.

b) For the variants that showed up at least four times in our sampling, we constructed rarefaction curves showing the cumulative count of those variants when samples were ordered by collection date. Here, we did not have to consider if two samples might have different variants at a given position, as that scenario did not occur after we only considered uncommon and common variants. We color coded the rarefaction curves based on if the variant was common, wave-1, wave-2, or wave-1+2. We labeled each line by variant position using a Matplotlib extension called matplotlib-label-lines.

c) This table simply shows the data in the previous figures in text format for the more frequent variants.

Figure 4

This tree was visualized using iTOL. See above for how the local phylogenetic tree was constructed.

**Supplemental figures**

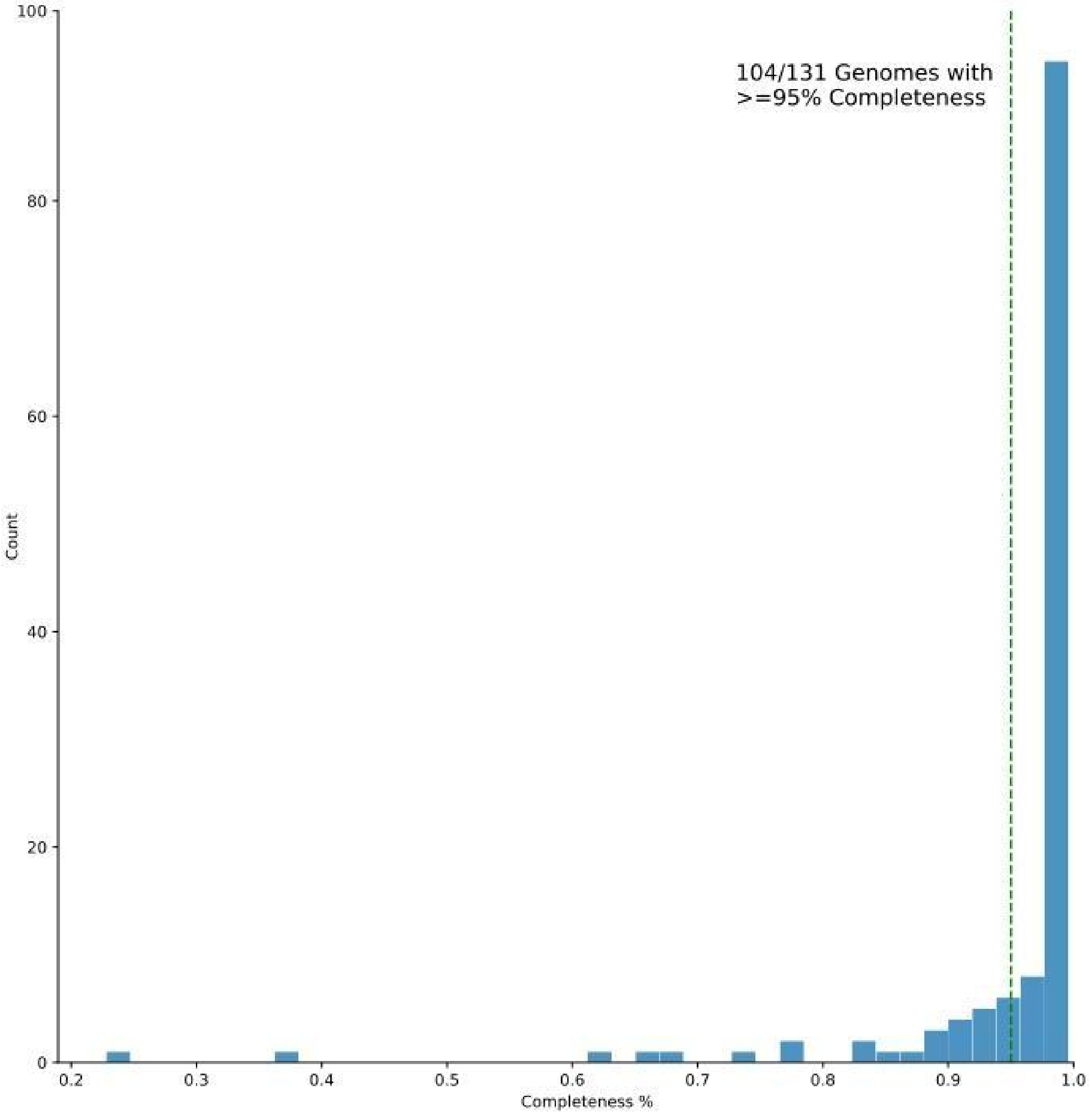


**Figure S1**. Histogram showing the distribution of completeness for all 131 genomes sequenced for this study.

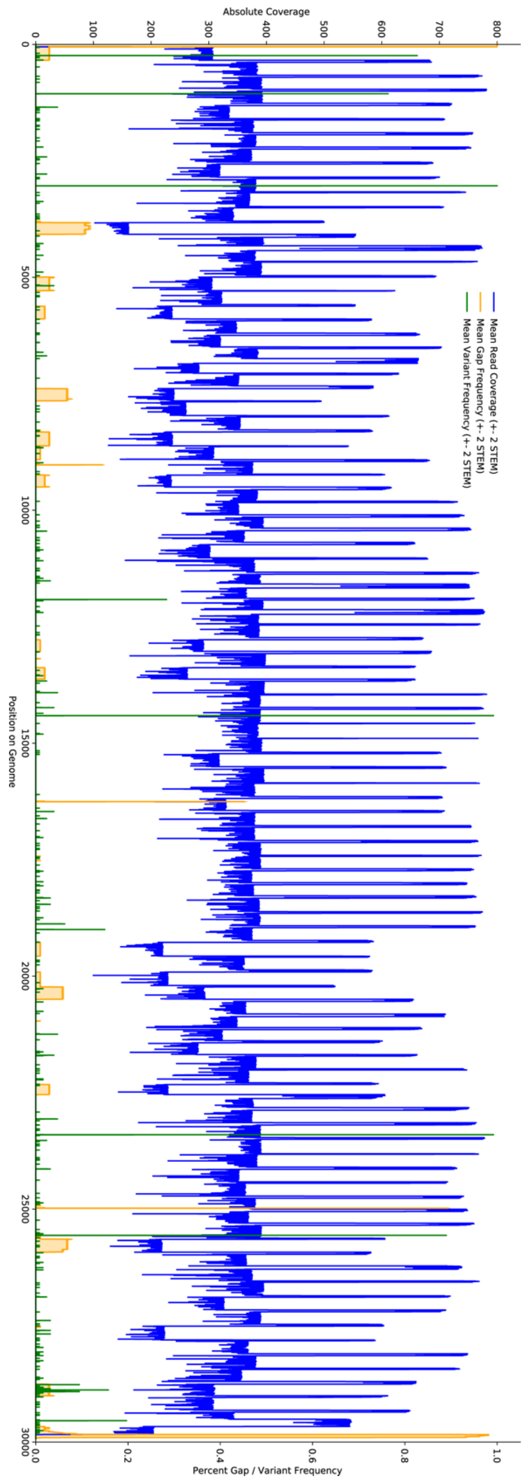


**Figure S2.** Visualization of coverage and gaps across all 104 included samples. In blue is the average coverage level at a given position. In yellow is the average frequency of gaps present at a given position. In green is the average frequency of variants at a given position.

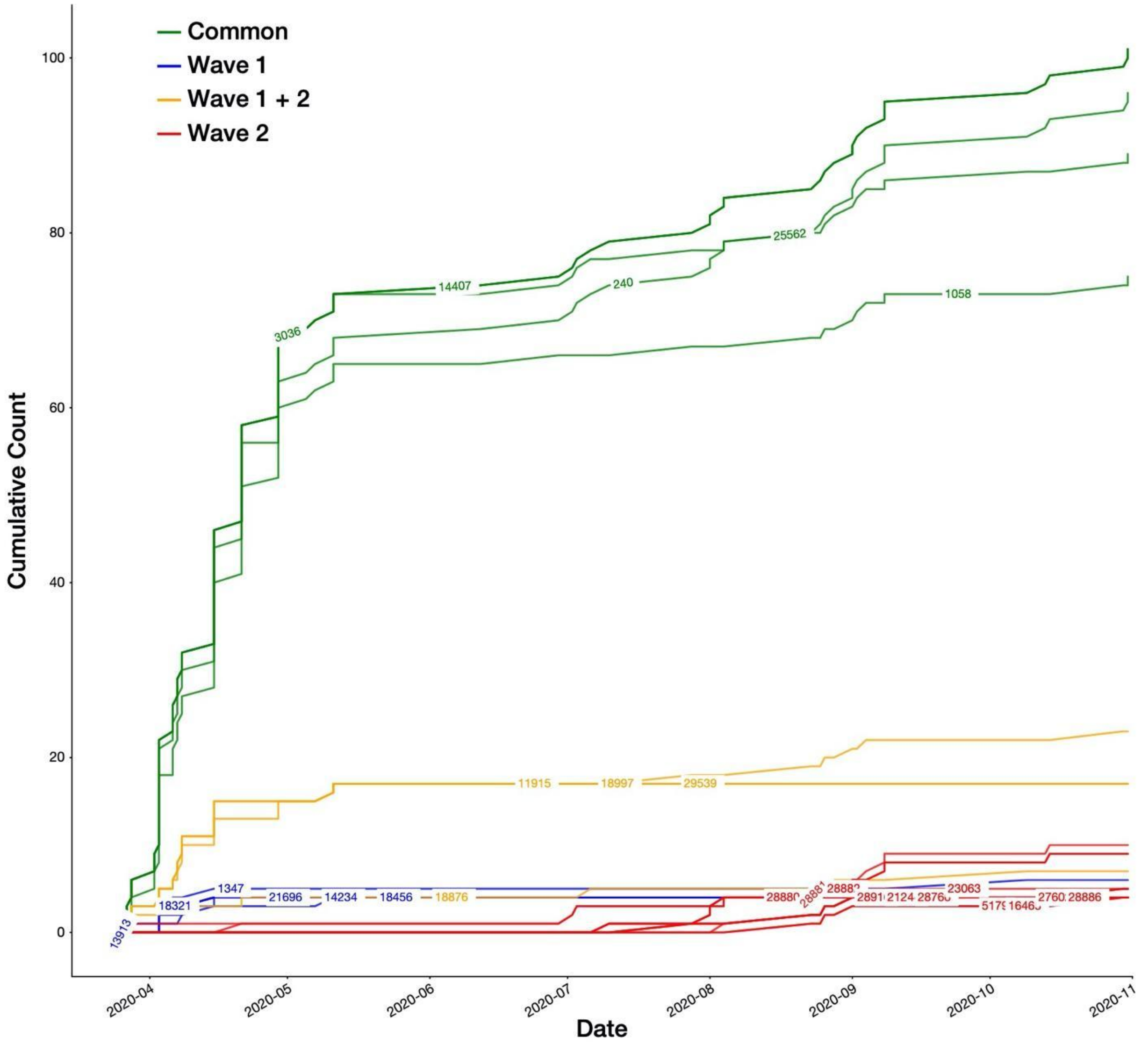


**Figure S3.** Distribution of cumulative counts of common and uncommon variants in the dataset. In contrast to Fig. 3B, the x-axis is ordered by date in order to demonstrate the temporal dynamics of variants in the Bronx.

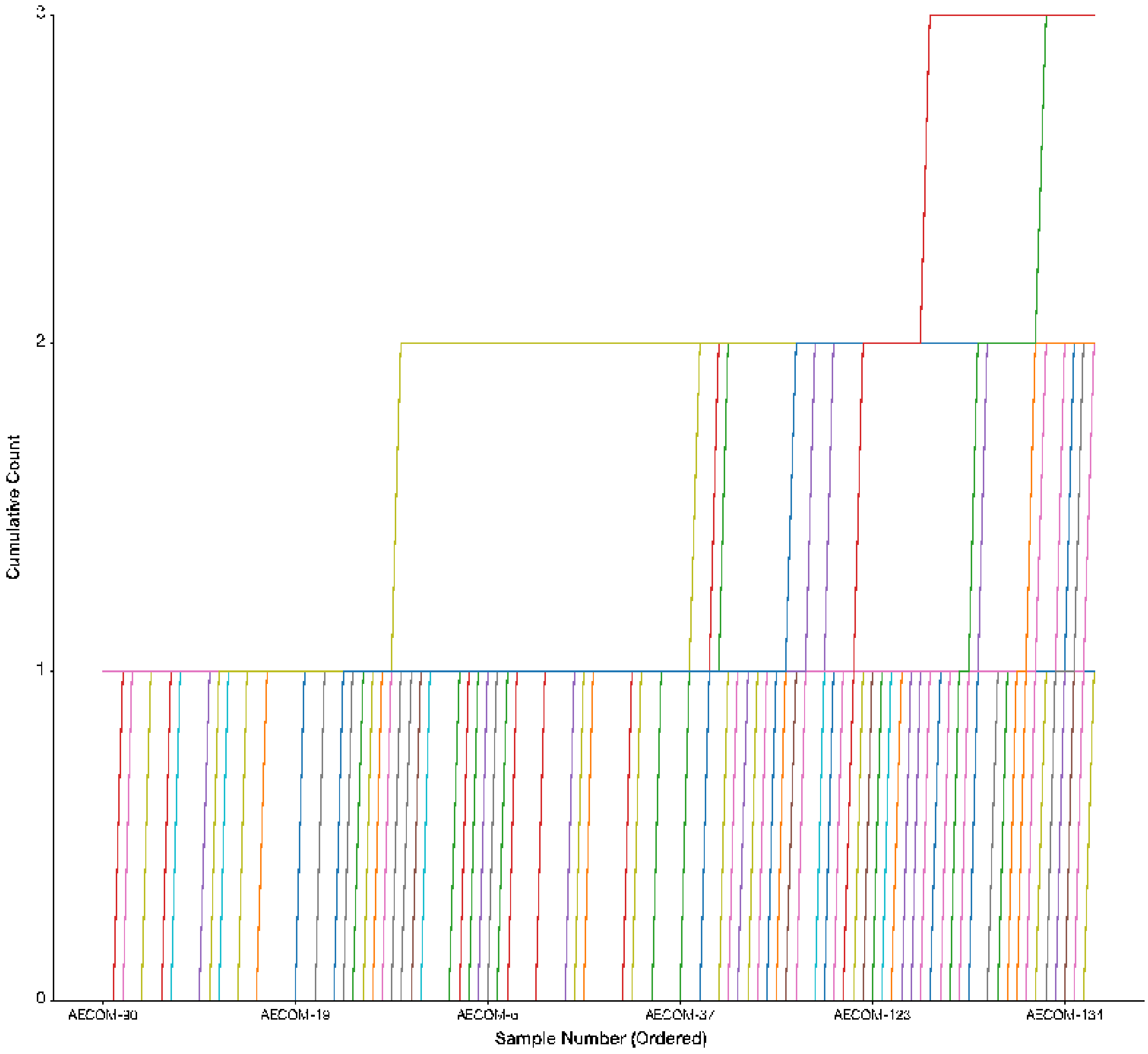


**Figure S4.** Rarefaction curve showing the cumulative counts of variants present less than four times total in the dataset. Samples are ordered by sample date. These rare variants were identified throughout the sampling period and do not seem to have a bias towards any particular time.

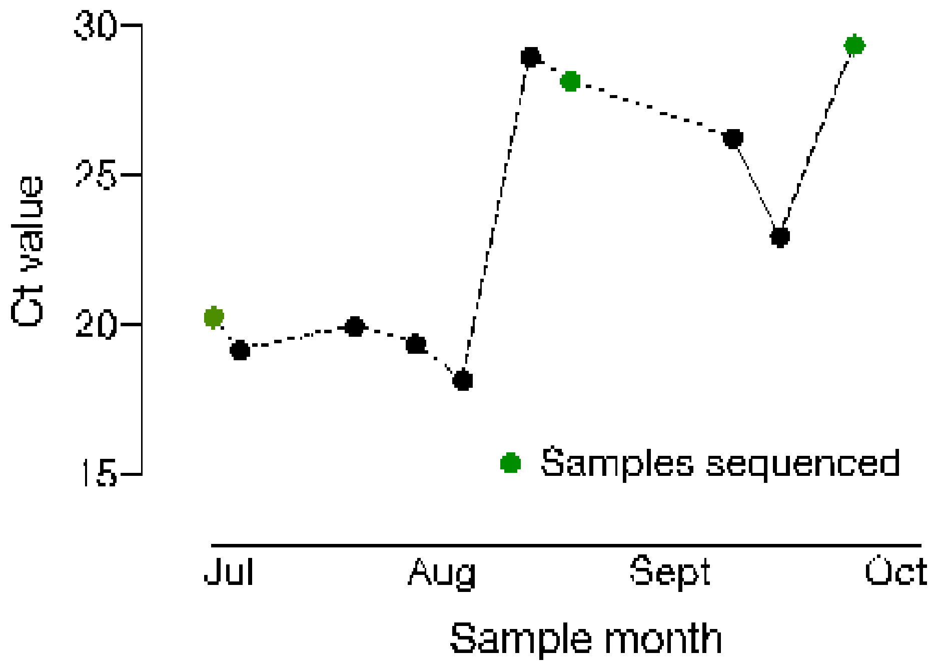


**Figure S5.** RT-PCR cycle thresholds of samples from Case 2 (see Table S1). Sequenced samples are highlighted in green.

| | Case 1 | Case 2 |
|---|---|---|
| Age | 12- 18 yrs | 12- 18 yrs |
| Sex | Female | Female |
| Comorbidities | No | Yes, see text |
| Date of first positive PCR (cycle time) | June 2020 (30.6) | July 2020 (20.3) |
| Symptoms at first presentation | Fever, sore throat, anosmia, ageusia | Lip ulcer, neutropenia |
| Duration of first illness | 6 days | Undefined (see text) |
| Date of next positive PCR (cycle threshold) | August 2020 (34.5) | Jul PCR1(19.2)<br>Jul PCR2 (20.0)<br>Aug PCR1 (19.4)<br>Aug PCR2 (18.2)<br>Aug PCR3 (29)<br>Aug PCR4 (28.2)<br>Sept PCR1 (26.3)<br>Sept PCR2 (23)<br>Oct PCR 1 (29.4) |
| Symptoms at second presentation | Fever, abdominal pain, diarrhea, myalgia | See text |
| Admission (Y/N) | N | Y |

**Table S1.** Patient characteristics of the re-infection (Case 1) and persistent infection (Case 2) cases.
